# Validation of a 0/2-hour high sensitivity cardiac troponin algorithm for suspected acute coronary syndrome in the emergency department

**DOI:** 10.1101/2025.05.15.25327732

**Authors:** Kaiser Permanente CREST Network Investigators, Dustin G. Mark, Jie Huang, Keane K. Lee, Dana R. Sax, Dustin W. Ballard, David R. Vinson, Mary E. Reed

## Abstract

**Background:** We implemented a high-sensitivity cardiac troponin I (hs-cTnI)-based algorithm for emergency department (ED) evaluation of possible non-ST elevation acute coronary syndromes (NSTE-ACS) within an integrated health system (The Kaiser Permanente Northern California [KPNC] NSTE-ACS algorithm).

**Methods:** Retrospective study of adult (18+ years) ED encounters for chest pain/discomfort with hs-cTnI testing (Access hsTnI, Beckman) at 21 KPNC medical centers between January 1, 2023 and June 30, 2024. Exclusion criteria were ST-elevation myocardial infarction (MI), leaving the ED against medical advice, lack of active KP health plan coverage, or an included encounter within 30 days prior. The primary outcome was 30-day MI or death. Sensitivity, specificity, negative predictive value (NPV), positive predictive value (PPV), and likelihood ratios (LR) were reported, with subgroup analyses by age, sex, coronary artery disease (CAD), chronic kidney disease (CKD), and ED disposition.

**Results:** There were 104,025 encounters in the final study cohort. Median age was 59 years, 45% were male, 18% had CAD, and 13% had CKD. The primary outcome occurred in 5.5% of encounters. Rule-out criteria were present in 70% of encounters with a sensitivity of 95.4% (95% CI: 94.8-96.0%), a NPV of 99.7% (95% CI: 99.6-99.7%) and an LR-of 0.05, while 7% of encounters met rule-in criteria with a specificity of 96.7% (95% CI: 96.6-96.8%), a PPV of 60.2% (95% CI: 59.3-61.1%) and an LR+ of 24.4. In subgroup analyses, rule-out criteria NPV was statistically below 99% in stage 4+ CKD (96.1%; 95% CI: 94.6-97.6%) and ischemic CAD (98.6%; 95% CI: 98.3-98.9%), though not among those selected for ED discharge (98.4%; 95% CI: 96.7-99.2% and 99.1%; 95% CI: 98.8-99.4%, respectively).

**Conclusions:** The KPNC NSTE-ACS evaluation algorithm demonstrated excellent overall performance. NPV was modestly diminished in ischemic CAD or advanced CKD, but this excess risk was largely mitigated by ED discharge disposition decisions.

## Introduction

Chest pain is the second most common presenting complaint during emergency department (ED) encounters, accounting for upwards of 5% of all ED visits in the United States.^1^ Efficient, efficacious, and safe risk stratification of patients with possible non-ST elevation acute coronary syndromes (NSTE-ACS) is, accordingly, an important clinical and operational goal. Progressive improvements in cardiac biomarker assay performance have been paramount in this regard, with high-sensitivity cardiac troponin (hs-cTn) assays representing the current state-of-the-art. Studies have demonstrated that hs-cTn-only algorithms, such as the European Society of Cardiology (ESC) 0/1 and 0/2 hour protocols, which stratify patients into low, intermediate and high risk classifications, can safely improve ED throughput and resource utilization.^2–6^ The High-Sensitivity Troponin in the Evaluation of Patients With Acute Coronary Syndrome (HIGH-STEACS) pathway is an endorsed alternative to the 0/1- or 0/2-hour ESC protocols that may increase overall rule-out efficacy in comparison, but has not been widely validated across various hs-cTn platforms.^7–13^

In advance of implementing an hs-cTnI assay (Access hsTnI, Beckman-Coulter, Brea, CA), and wanting to combine features of both the 0/2-hour ESC protocol and HIGH-STEACS pathway, we designed a hybrid 0/2-hour algorithm (the Kaiser Permanente Northern California [KPNC] NSTE-ACS algorithm). We previously reported a preliminary validation of the KPNC NSTE-ACS algorithm which demonstrated accuracy and safety in terms of 30-day MI incidence as well a greater proportion of patients ruled-out as compared to the 0/2-hour ESC protocol.^14^ The goal of the current study is to present a larger validation of the KPNC NSTE-ACS algorithm, extend the safety assessment to include 30-day all-cause death, and explore differential risk among clinical subgroups, particularly known CAD given prior reports that negative predictive values (NPV) can drop below a 99% safety threshold when applying the ESC 0/1- or 0/2-hour protocol to this subgroup.^15, 16^

## Methods

### Study design and setting

We performed a retrospective cohort study of adult (aged 18 years and older) ED patient encounters within KPNC between January 1, 2023 and June 30, 2024. KPNC operates 21 community medical center-based EDs serving a diverse population with over 1.5 million annual ED visits. KPNC health plan members include approximately 33% of the population in areas served and are representative of the demographic and socioeconomic diversity of the surrounding and statewide population.^17^ All EDs are staffed exclusively by board-certified emergency physicians.

Patients were included if they had a chief complaint of chest pain or discomfort and hs-cTn testing within six hours of ED arrival. Patients were excluded if they had an ED diagnosis of ST-elevation MI, an ED disposition of left against medical advice, lack of active KPNC health plan coverage at the index visit and during the following month (for completeness of outcome capture), or if there was a study eligible encounter within 30 days prior. Reporting adhered to the Standards for Reporting Diagnostic accuracy studies (STARD) guidelines. The KPNC Institutional Review Board approved the study with a waiver of informed consent. Requests to access the data set from qualified researchers trained in human subject confidentiality protocols may be sent to KPNC at.

### Hs-cTnI assay and KPNC NSTE-ACS algorithm implementation

KPNC introduced the Access hsTnI assay across all facility laboratories on November 16, 2022. Lithium heparin plasma samples were run on the Beckman Access 2 immunoassay system, which has a reported assay limit of quantitation (20% coefficient of variation, CV) of 2.0 ng/L, a 10% CV of 4.1 ng/L, and an estimated maximal standard deviation of 0.89 ng/L at a mean concentration of 8.9 ng/L (total assay imprecision).^18^ Sex-specific 99^th^ centile upper reference limits (12 ng/L for females, 20 ng/L for males) were adopted per manufacturer recommendations. At the same time, an hs-cTnI-based algorithm was introduced for the risk stratification of patients with possible non-ST elevation acute coronary syndrome (the KPNC NSTE-ACS algorithm).

The KPNC NSTE-ACS algorithm was designed as a hybrid of the HIGH-STEACS pathway rule-out criteria (Supplemental Figure 1) and the ESC 0/2-hour protocol rule-in criteria (Supplemental Figure 2), as described in Figure 1, and places patients into one of four risk strata (very low risk, low risk, indeterminate risk, or high risk). Risk-based recommendations were as follows: 1) expedited ED discharge for very low risk and low risk encounters (rule-out); 2) serial troponin testing and consideration of urgent stress testing for indeterminate risk encounters (observation), 3) admission/consultation for high risk encounters (rule-in). Of note, presence of an ischemic-appearing electrocardiogram (ECG) was a disqualification for expedited ED discharge and resulted in a minimum designation of indeterminate risk.^8, 19^

**Figure 1.**
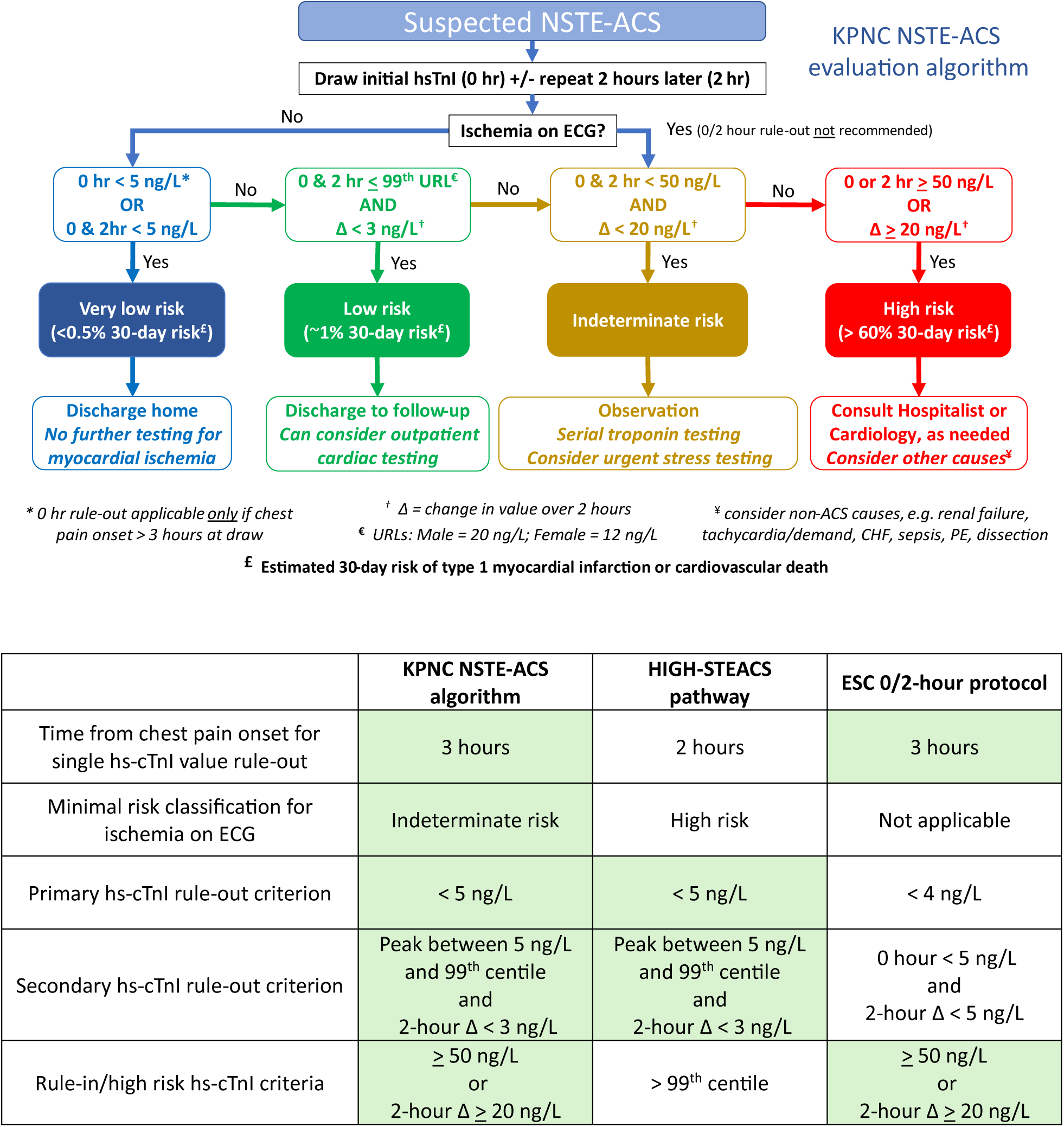
KPNC NSTE-ACS evaluation algorithm. The KPNC NSTE-ACS evaluation algorithm was available for reference as shown, including risk strata assignment (very low, low, indeterminate or high) with corresponding disposition and management recommendations. Hs-cTnI results were obtained using the Access hsTnI assay (Beckman-Coulter). Key shared features between the KPNC NSTE-ACS algorithm and the parent algorithms (HIGH-STEACS pathway and ESC 0/2-hour protocol) are shown in a separate table, highlighted in green. Abbreviations: ECG = electrocardiogram; ESC = European Society of Cardiology; Hs-cTnI = high-sensitivity cardiac troponin I; HIGH-STEACS = High-Sensitivity Troponin in the Evaluation of Patients With Acute Coronary Syndrome; KPNC = Kaiser Permanente Northern California; LR = likelihood ratio; MI = myocardial infarction; NPV = negative predictive value; NSTE-ACS = non-ST elevation acute coronary syndrome; PPV = positive predictive value; URL = upper reference limit (i.e. 99^th^ centile)

Very low risk designations (principal rule-out criterion) were based on an hs-cTnI value below 5 ng/L being obtained a minimum of three hours from chest pain onset, consistent with American College of Cardiology expert consensus guidance, and allowed for a single-troponin rule-out.^8^ Low risk designations (secondary rule-out criterion) were made if the highest of the 0- and 2-hour hs-cTnI values was between 5 ng/L and the 99^th^ centile and there was a 0/2-hour difference (“delta”) in hs-cTnI values of less than 3 ng/L. This delta was chosen to approximate two standard deviations of the total assay imprecision estimate. High risk designations (rule-in criterion) were an hs-cTnI value of ≥ 50 ng/L or a 0/2-hour delta of ≥ 20 ng/L.^6^ Remaining hs-cTnI profiles were classified as indeterminate risk. For the purposes of this study, patients with only one hs-cTnI measurement within six hours of ED arrival and that resulted between 5 and 49 ng/L are reported separately from the indeterminate risk group as having “insufficient data” for risk designation.

Educational sessions were provided to all KPNC emergency physicians prior to implementation, and clinical reference weblinks to the algorithm were available within the electronic health record (EHR; Epic, Verona, WI). In addition, electronic clinical decision support (eCDS) for algorithm interpretation was available to emergency physicians via a web-based platform (RISTRA) embedded within the EHR, which also allowed for prospective confirmation of algorithm reference during a given clinical encounter.^20^ Notably, there was no specific prompt to use the RISTRA eCDS outside of the educational sessions, and thus all RISTRA eCDS use was clinician-initiated.

### Variables

Demographics and comorbidities were electronically extracted from the EHR. Comorbidities were determined using International Classification of Diseases, 10^th^ Revision (ICD-10) diagnoses within the active problem list at the time of the index encounter, including resolved problems when indicated (Supplemental Methods). Presence of ischemic-appearing ECGs was determined using a validated text string analysis methodology of the final ECG interpretation, as previously described (Supplemental Methods).^21^

### Outcomes

The primary outcome was a composite of 30-day MI or death, inclusive of index events. Secondary outcomes were ED disposition (discharge, observation or hospital admission), 30-day non-invasive (functional or anatomic) or invasive coronary testing, and 30-day coronary revascularization (percutaneous or surgical). Death was determined using a composite death database which draws from internal KPNC mortality records, the California Department of Public Health Vital Records, and the Social Security Death Index. Procedural outcomes and coronary tests were determined using ICD-10 or current procedural terminology diagnostic code. MI outcomes were ascertained using both automatic criteria and adjudication by manual chart review. Automatic criteria for MI were an internal ICD-10 diagnostic code for non-ST-elevation MI or ST-elevation MI (Supplemental Methods) along with at least one hs-cTnI value above the 99^th^ centile within the 30-day outcome window (in accordance with the fourth universal definition of MI^22^), or any corresponding ICD-10 diagnostic code within an outside claims database (no laboratory confirmation required). Adjudication of MI by manual chart review was conducted if there was an internal ICD-10 diagnostic code for MI without an hs-cTnI value above the 99^th^ centile within the 30-day outcome window, or if there was both an hs-cTnI value above the 99^th^ centile and an ICD-10 diagnosis of acute coronary syndrome, type 2 MI or unstable angina within 30 days following a very low risk or low risk encounter (to better ensure algorithm rule-out safety). Manual review was independently performed by two emergency physicians using all available clinical notes, cardiac studies and ECGs within the EHR to determine the presence or absence of a type 1 MI, as defined by the fourth universal definition of MI, with disagreements adjudicated by a cardiologist. Details on ICD-10 and procedural codes are available in the Supplemental Methods.

### Data analysis

Outcome incidence was reported for each risk strata, and test characteristics were reported for the primary outcome and its components, including sensitivity, specificity, negative predictive value (NPV), positive predictive value (PPV), and likelihood ratio (LR), including negative LR (LR-) or positive LR (LR+) where applicable. Algorithm overall rule-out performance was assessed using the combination of the principal (very low risk strata) and secondary (low risk strata) rule-out criterion, while algorithm rule-in performance was assessed using the high risk strata. Additionally, test characteristics for the principal and secondary rule-out criterion were assessed individually. Principal rule-out criterion test characteristics were calculated inclusive of encounters with insufficient data, since those encounters would not have been classified as very low risk (i.e. hs-cTnI was not < 5 ng/L). For secondary rule-out criterion test characteristic calculations, to allow for a binary test definition and better represent clinical decision making in that scenario, encounters designated as very low risk were not included.

Subgroup analyses of rule-out and rule-in test characteristics for the primary outcome were performed according to age (< 45, 45-64, ≥ 65), sex, history of chronic kidney disease (CKD), and known coronary artery disease (CAD). CKD was further categorized as stage 4+ (estimated glomerular filtration rate < 30 mL/min/1.73m^2^), stage 3 (estimated glomerular filtration rate 30-59 mL/min/1.73m^2^), or absent. CAD was categorized as either ischemic (prior coronary revascularization or MI), nonischemic, or absent. To further characterize independent risks by subgroup, and to account for correlations due to repeat patient encounters, we modeled the primary outcome (dependent variable) within the rule-out and rule-in groups separately using a generalized estimating equation (binomial family and logit link) controlling for the above subgroups in addition to diabetes, smoking, obesity, peripheral artery disease, cerebrovascular disease, and facility, reported as adjusted odds ratios (aOR). Finally, to assess the performance of the algorithm in clinical practice, we examined the incidence of outcomes within risk strata according to final ED disposition (discharge vs observation or admission), reporting outcome differences as relative risks (RR).

Outcome incidences and test characteristics were calculated with Wilson 95% confidence intervals. Differences in proportions were assessed using Chi-squared or Fisher’s exact tests. P-values <0.05 were considered statistically significant. Analyses were performed with Stata 17.0 (StataCorp, College Station, TX).

### Sensitivity analysis

Sensitivity analyses for algorithm performance and test characteristics were performed on the subset of encounters in which RISTRA eCDS was accessed by clinicians to aid in KPNC NSTE-ACS algorithm interpretation. As noted above, clinicians accessed RISTRA eCDS without any real-time prompts, and thus this subset of encounters might be considered as having a more specific indicator of clinical concern for possible ACS as compared to the entire study cohort.

## Results

There were 104,025 encounters among 93,433 patients in the study cohort (Figure 2). The median age was 59 years, 55% were female sex, 25% had diabetes, 13% had ischemic CAD, and 3% had stage 4+ CKD (Table 1). A primary outcome of 30-day MI or death occurred in 5.5% of encounters (30-day MI in 5.0% and death in 0.7%), with 81% of encounters resulting in discharge directly from the ED (Table 2). Non-invasive coronary testing, invasive coronary testing, and coronary revascularization were performed within 30 days in 19%, 6.0%, 2.9% of encounters, respectively. The incidence of the primary outcome by risk strata (Table 2 and Supplemental Figure 3) was as follows: very low-risk (0.1%; 95% CI: 0.1-0.2%), low-risk (1.2%; 95% CI: 1.1-1.4%), indeterminate-risk (6.6%; 95% CI: 6.2-7.1%) and high-risk (60.2%; 95% CI: 59.3-61.1%) with corresponding LRs of 0.02, 0.20, 1.14 and 24.3, respectively.

**Figure 2.**
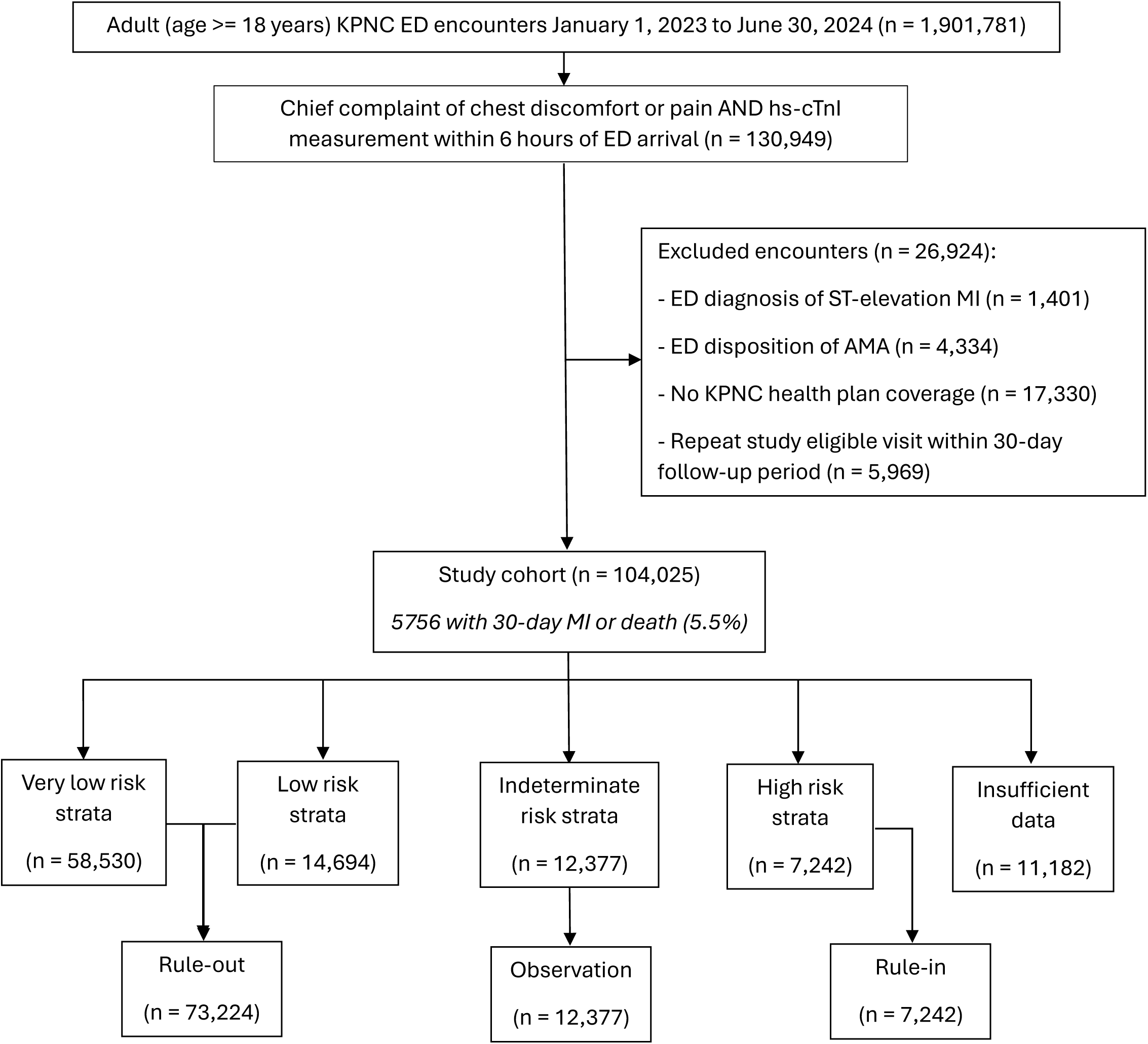
Study flow diagram. Summary of 104,025 study encounters, with distribution by risk strata (very low, low, indeterminate, high) and summary disposition recommendation (rule-out, observation, rule-in) according to the KPNC NSTE-ACS evaluation algorithm. Note that 11,182 encounters had insufficient troponin data to allow for definitive risk strata assignment, though were at least excluded from the very low risk strata (i.e. hs-cTnI value not < 5 ng/L). Abbreviations: AMA = against medical advice; ED = emergency department; Hs-cTnI = high-sensitivity cardiac troponin I; KPNC = Kaiser Permanente Northern California; NSTE-ACS = non-ST elevation acute coronary syndrome.

**Table 1.** Study cohort demographics and characteristics

Validation of the KPNC NSTEMI-ACS algorithm
|  |  | Risk strata |  |  |  |  |
| --- | --- | --- | --- | --- | --- | --- |
| N (%) | Overall<br>n = 104025 | Very low<br>n = 58530 | Low<br>n = 14694 | Indeterminate<br>n = 12377 | High<br>n = 7242 | Insufficient<br>data<br>n = 11182 |
| Age, median years (IQR) | 59 (44-71) | 50 (39-62) | 69 (58-78) | 70 (57-80) | 69 (58-79) | 68 (55-78) |
| < 45 | 26179 (25) | 21820 (37) | 1173 (8) | 1243 (10) | 567 (8) | 1376 (12) |
| 45-64 | 38341 (37) | 24537 (42) | 4558 (31) | 3514 (28) | 2269 (31) | 3463 (31) |
| ≥ 65 | 39505 (38) | 12173 (21) | 8963 (61) | 7620 (62) | 4406 (61) | 6343 (57) |
| Female, n | 57580 (55) | 36518 (62) | 6227 (42) | 6710 (54) | 2792 (39) | 5333 (48) |
| Race/ethnicity |  |  |  |  |  |  |
| White | 46209 (44) | 23572 (40) | 7548 (51) | 6066 (49) | 3689 (51) | 5334 (48) |
| Black | 12157 (12) | 6422 (11) | 1716 (12) | 1660 (13) | 809 (11) | 1550 (14) |
| Hispanic | 25253 (24) | 16594 (28) | 2784 (19) | 2272 (18) | 1306 (18) | 2297 (21) |
| Asian | 18186 (17) | 10690 (18) | 2330 (16) | 2102 (17) | 1296 (18) | 1768 (16) |
| Other | 2220 (2) | 1252 (2) | 316 (2) | 277 (2) | 142 (2) | 233 (2) |
| Diabetes | 26202 (25) | 9936 (17) | 5095 (35) | 4569 (37) | 2980 (41) | 3622 (32) |
| Hypertension | 31910 (31) | 9174 (16) | 7256 (49) | 6505 (53) | 3804 (53) | 5171 (46) |
| Hypercholesterolemia, | 47144 (45) | 18890 (32) | 9137 (62) | 7766 (63) | 4866 (67) | 6485 (58) |
| Obesity (BMI ≥ 30) | 44340 (43) | 25552 (44) | 6308 (43) | 4875 (39) | 2788 (38) | 4817 (43) |
| Family history of<br>premature CAD | 5919 (6) | 3308 (6) | 829 (6) | 689 (6) | 517 (7) | 576 (5) |
| Smoking | 8259 (8) | 4541 (8) | 1105 (8) | 976 (8) | 695 (10) | 942 (8) |
| Peripheral artery disease | 2267 (2) | 310 (1) | 569 (4) | 580 (5) | 434 (6) | 374 (3) |
| Cerebrovascular disease | 8365 (8) | 2120 (4) | 1967 (13) | 1916 (15) | 1032 (14) | 1330 (12) |
| Chronic renal disease |  |  |  |  |  |  |
| Stage 3 | 10444 (10) | 1980 (3) | 2689 (18) | 2503 (20) | 1404 (19) | 1868 (17) |
| Stage 4+ | 3129 (3) | 83 (0) | 559 (4) | 1094 (9) | 874 (12) | 519 (5) |
| Coronary artery disease |  |  |  |  |  |  |
| Nonischemic | 5650 (5) | 1523 (3) | 1299 (9) | 1247 (10) | 686 (9) | 895 (8) |
| Ischemic | 13981 (13) | 2710 (5) | 3392 (23) | 3363 (27) | 2461 (34) | 2055 (18) |
| Ischemic ECG | 4338 (4) | 0 (0) | 0 (0) | 2670 (22) | 1136 (16) | 532 (5) |
| Time from ED arrival to<br>first troponin result,<br>minutes (IQR) | 100 (79-132) | 102 (80-134) | 95 (76-122) | 97 (78-126) | 99 (78-130) | 106 (82-145) |
| Time between first and<br>second troponin results,<br>minutes (IQR) | 124 (105-148) | 123 (104-145) | 128 (111-152) | 121 (98-146) | 125 (103-156) | NA |
| Peak troponin within 6<br>hours, ng/L, median<br>(IQR) | 4 (2-8) | 2 (0-3) | 7 (6-10) | 15 (8-25) | 183 (77-759) | 7 (5-10) |

Abbreviations: BMI = body mass index; CAD = coronary artery disease; ECG = electrocardiogram; HEART = History, Electrocardiogram, Age, Risk factors, Troponin.

**Table 2.** Primary and secondary outcomes.

Validation of the KPNC NSTE-ACS algorithm
|  |  | Risk strata |  |  |  |  |
| --- | --- | --- | --- | --- | --- | --- |
| N (%) | Overall, n = 104025 | Very low n = 58530 | Low n = 14694 | Indeterminate n = 12377 | High n = 7242 | Insufficient data n = 11182 |
| 30-day MI or death | 5756 (5.5) | 69 (0.1) | 180 (1.2) | 817 (6.6) | 4358 (60) | 332 (3.0) |
| 30-day MI | 5158 (5.0) | 36 (0.1) | 85 (0.6) | 645 (5.2) | 4209 (58) | 183 (1.6) |
| 30-day death | 763 (0.7) | 33 (0.1) | 100 (0.7) | 189 (1.5) | 286 (3.9) | 155 (1.4) |
| ED disposition |  |  |  |  |  |  |
| Discharge | 84446 (81) | 55521 (95) | 11783 (80) | 7303 (59) | 767 (11) | 9072 (81) |
| Observation | 9269 (9) | 1860 (3) | 1687 (12) | 2760 (22) | 2042 (28) | 920 (8) |
| Admission | 10310 (10) | 1149 (2) | 1224 (8) | 2314 (19) | 4433 (61) | 1190 (11) |
| 30-day testing |  |  |  |  |  |  |
| 30-day non-invasive coronary testing | 19354 (19) | 8595 (15) | 3723 (25) | 3756 (30) | 1609 (22) | 1671 (15) |
| 30-day invasive coronary testing | 6245 (6.0) | 411 (0.7) | 495 (3.4) | 1032 (8.3) | 3914 (54) | 393 (3.5) |
| 30-day coronary revascularization |  |  |  |  |  |  |
| Any | 2973 (2.9) | 107 (0.2) | 154 (1.1) | 419 (3.4) | 2127 (29) | 166 (1.5) |
| PCI | 2164 (2.1) | 74 (0.1) | 103 (0.7) | 285 (2.3) | 1589 (22) | 113 (1.0) |
| CABG | 855 (0.8) | 34 (0.1) | 55 (0.4) | 142 (1.2) | 566 (7.8) | 58 (0.5) |

Abbreviations: CABG = coronary artery bypass graft; ED = emergency department; MI = myocardial infarction PCI = percutaneous coronary intervention.

### Rule-out criteria performance

Rule-out criteria were met in 73224 (70%) encounters with a sensitivity of 95.4% (95% CI: 94.8-96.0%), a specificity of 83.5% (95% CI: 83.2-83.7%), an LR-of 0.05 (95% CI: 0.05-0.06), and a NPV of 99.7% (95% CI: 99.6-99.7%, Table 3 and Supplemental Figure 3). Sensitivity and NPV for the primary outcome were marginally higher for the principal rule-out criterion with a sensitivity of 98.8% (95% CI: 98.5-99.1%) and an NPV of 99.9% (95% CI: 99.9-99.9%) as compared to the secondary rule-out criterion, which had a sensitivity of 96.6% (95% CI: 96.1-97.1%) and an NPV of 98.8% (95% CI: 98.6-98.9%), reflecting a significant difference in rule-out performance between the two criterion. Further test characteristics for the primary outcome and its individual components are presented in Table 3.

**Table 3.** Test characteristics of rule-out criteria and constituent criterions for the primary outcome and components.

|  |  | TP | FN | FP | TN | Sensitivity | Specificity | NPV | LR- |
| --- | --- | --- | --- | --- | --- | --- | --- | --- | --- |
| Rule-out criteria<br>n = 73224 *<br>(70% efficacy) | 30-day MI or death | 5175 | 249 | 1444<br>4 | 7297<br>5 | 95.4<br>(94.8-96.0) | 83.5<br>(83.2-83.7) | 99.7<br>(99.6-99.7) | 0.05<br>(0.05-0.06) |
|  | 30-day MI | 4854 | 121 | 1476<br>5 | 7310<br>3 | 97.6<br>(97.1-98.0) | 83.2<br>(83.0-83.4) | 99.8<br>(99.8-99.9) | 0.03<br>(0.02-0.03) |
|  | 30-day death | 475 | 133 | 1914<br>4 | 7309<br>1 | 78.1<br>(74.6-81.4) | 79.2<br>(79.0-79.5) | 99.8<br>(99.8-99.8) | 0.28<br>(0.24-0.32) |
| Primary rule-out criterion<br>n = 58530 **<br>(56% efficacy) | 30-day MI or death | 5687 | 69 | 3980<br>8 | 5846<br>1 | 98.8<br>(98.5-99.1) | 59.5<br>(59.2-59.8) | 99.9<br>(99.9-99.9) | 0.02<br>(0.02-0.03) |
|  | 30-day MI | 5122 | 36 | 4037<br>3 | 5849<br>4 | 99.3<br>(99.1-99.5) | 59.2<br>(58.9-59.5) | 99.9<br>(99.9-100) | 0.01<br>(0.01-0.02) |
|  | 30-day death | 730 | 33 | 4476<br>5 | 5849<br>7 | 95.7<br>(94.0-97.0) | 56.7<br>(56.4-57.0) | 99.9<br>(99.9-100) | 0.08<br>(0.05-0.11) |
| Secondary rule-out criterion<br>n = 14694 ***<br>(14% efficacy) | 30-day MI or death | 5175 | 180 | 1444<br>4 | 1451<br>4 | 96.6<br>(96.1-97.1) | 50.1<br>(49.5-50.7) | 98.8<br>(98.6-98.9) | 0.07<br>(0.06-0.08) |
|  | 30-day MI | 4854 | 85 | 1476<br>5 | 1460<br>9 | 98.3<br>(97.9-98.6) | 49.7<br>(49.2-50.3) | 99.4<br>(99.3-99.5) | 0.03<br>(0.03-0.04) |
|  | 30-day death | 475 | 100 | 1914<br>4 | 1459<br>4 | 82.6<br>(79.3-85.6) | 50.0<br>(49.5-50.5) | 99.5<br>(99.4-99.6) | 0.35<br>(0.29-0.42) |

\* Test characteristics for the rule-out criteria were calculated after excluding encounters with insufficient troponin data for risk category assignment (n = 11182). Reference cohort n = 92843.
\*\* Test characteristics for the primary rule-out criterion (hs-cTnI < 5 ng/L, very low risk strata) were calculated with inclusion of encounters with insufficient troponin data (n = 11182) since all such encounters had a troponin value of at least 5 ng/L and thus would not have been considered very low risk. Reference cohort n = 104025.
\*\*\* To both allow for a binary test definition and characterize the incremental value of the secondary rule-out criterion (hs-cTnI 5 ng/L to 99<sup>th</sup> centile with a 2-hour delta < 3 ng/L), test characteristics were calculated after exclusion of encounters with insufficient troponin data (n = 11182) and encounters classified as very low risk (n = 58530). Reference cohort n = 34313.
Abbreviations: hs-cTnI = high-sensitivity cardiac troponin I; FN = false negative; FP = false positive; MI = myocardial infarction; LR- = negative likelihood ratio; NPV = negative predictive value; TN = true negative; TP = true positive.

### Subgroup analysis of rule-out criteria

Subgroup analysis of the rule-out criteria (Figure 3) revealed marginally lower NPV for male sex and more notable decreases in NPV with increasing age and degrees of CKD or CAD. The largest difference in NPV was encounters with stage 4+ CKD vs no CKD (96.1% vs 99.7%, absolute difference 3.8%; 95% CI: 2.5-5.7%, p <0.001) followed by ischemic CAD vs no CAD (98.6% vs 99.8%, absolute difference 1.2%; 95% CI: 0.9-1.5%, p <0.001). All decreases in NPV were attributable to both increases in outcome incidence and decreases in specificity, as sensitivities remained stable (range 94.4% to 96.1%, Supplemental Table 1). Separate examination of principal and secondary rule-out criterion revealed similar findings (Supplemental Figure 4 and Supplemental Figure 5). Multivariable analysis (Supplemental Table 2) showed that age ≥ 65 had strongest independent association with the primary outcome among rule-out encounters (aOR 7.2; 95% CI: 3.7-14.3) followed by stage 4+ CKD (aOR 5.2; 95% CI: 3.2-8.3), and ischemic CAD (aOR 2.2; 95% CI: 1.6-3.1).

**Figure 3.**
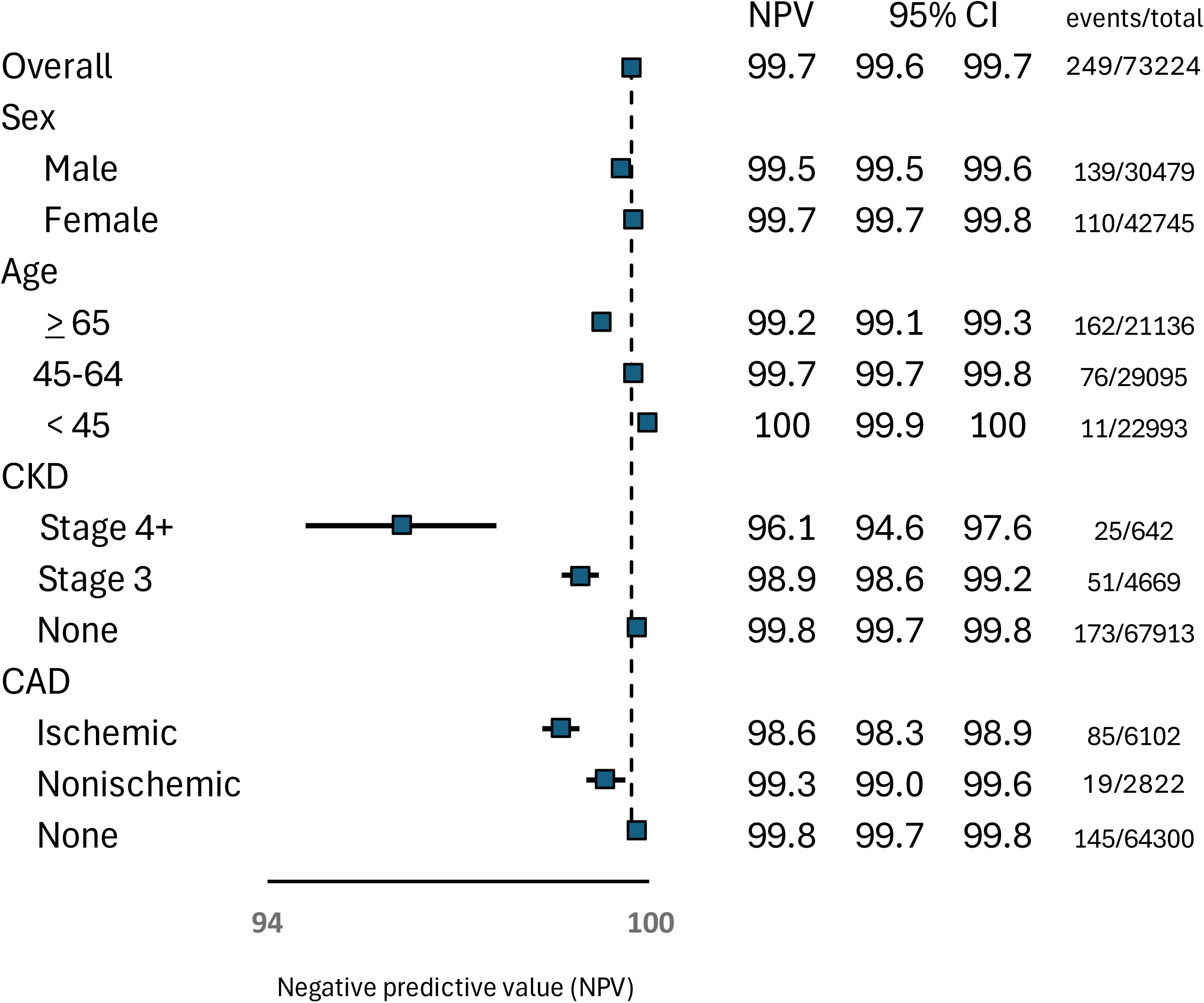
Subgroup analysis of rule-out encounters for 30-day myocardial infarction or death. Forest plot summary of rule-out criteria negative predictive value for 30-day myocardial infarction or death within key clinical subgroup strata (by age, sex, coronary artery disease, or chronic kidney disease). Abbreviations: CAD = coronary artery disease; CKD = chronic kidney disease; NPV = negative predictive value.

### Rule-in criteria performance

Rule-in criteria were met in 7242 (7%) of encounters with a primary outcome sensitivity of 80.4% (95% CI: 79.3-81.4%), specificity of 96.7% (95% CI: 96.6-96.8%), an LR+ of 24.4 (95% CI: 23.4-25.3), and a PPV of 60.2% (95% CI: 59.3-61.1%, Supplemental Table 3 and Supplemental Figure 3). Subgroup analysis (Supplemental Table 4 and Supplemental Figure 6) showed that PPV point estimates dropped below 50% for age < 45 (43.4%: 95% CI: 40.5-46.3%), stage 4+ CKD (45.8%; 95% CI: 43.5-48.1%) and nonischemic CAD (49.1%; 95% CI: 46.3-52.0%), The relative decreases in PPV among these subgroups was consistent in multivariate modeling (Supplemental Table 5).

### Performance in clinical practice (ED disposition)

Overall, 84446 (81%) of study encounters resulted in ED discharge, of which 450 (0.53%; 95% CI: 0.48-0.58%) experienced the primary outcome of 30-day MI or death (Supplemental Table 6). ED discharges varied in proportion by risk strata: very low risk (95%), low risk (80%), indeterminate risk (59%), high risk (11%), and insufficient data (81%). Of encounters meeting rule-out criteria, 67304/73224 (92%) were discharged directly from the ED. The incidence of the primary outcome in the rule-out group following ED discharge was 126/67304 (0.19%, Supplemental Table 7). Comparatively, among rule-out encounters resulting in observation or admission, the primary outcome occurred in 123/5920 (2.1%). Thus, selection of rule-out encounters for ED discharge effectively provided an added layer of risk stratification (RR 0.09; 95% CI: 0.7-0.11).

Similar degrees of risk discernment through selection for ED discharge were noted in subgroup analysis of rule-out encounters, specifically among those subgroups with the highest independent association with the primary outcome (Supplemental Table 7 and Supplemental Figure 7). For encounters with age ≥ 65, 18063/21136 (85%) were discharged from the ED with lower primary outcome events compared to encounters selected for observation or admission (72/18063 [0.39%] vs 90/3073 [2.93%], RR 0.13; 95% CI: 0.10-0.18); in CKD4+, 429/642 (67%) were discharged (primary outcome in 7/429 [1.6%] vs 18/213 [8.5%], RR 0.19; 95% CI: 0.08-0.46); and in known ischemic CAD, 4924/6102 (81%) were discharged (primary outcome in 43/4924 [0.87%] vs 42/1178 [3.6%], RR 0.24; 95% CI: 0.16-0.37).

### Sensitivity analysis (RISTRA eCDS use)

Demographics for encounters with clinician-initiated use of RISTRA eCDS (n = 3259) were comparable to the main cohort (Supplemental Table 8), though the primary outcome occurred less frequently (3.0% vs 5.5%) with a lower proportion of encounters meeting rule-in criteria (3% vs 7%, Supplemental Table 9 and Supplemental Figure 8), likely being reflective of the limited use of eCDS in overtly high-risk scenarios (e.g. markedly abnormal hs-cTnI values). Rule-out criteria performed similarly in the RISTRA eCDS encounters as compared to the main cohort (primary outcome sensitivity 95.7%; 95% CI: 89.6-98.3% and NPV 99.8%; 95% CI: 99.6-99.9%, Supplemental Figure 8 and Supplemental Table 10). Notably, NPV was higher for the secondary rule-out criterion (99.7%; 95% CI: 98.9-99.9%) as compared to the full study cohort, mostly attributable to a significantly lower incidence of death (0% vs 0.68%, p = 0.02).

## Discussion

### Rule-out criteria performance

Our findings support the use of the KPNC NSTE-ACS algorithm to safely identify patients at low risk for 30-day MI or death in US ED populations. The NPV for 30-day MI or death in our study (99.7%) is consistent with the early rule-out performance of the HIGH-STEACS pathway (from which the KPNC NSTE-ACS rule-out criteria were derived) when using an hs-cTnI assay (NPVs of 99.5% and 99.6%).^7, 10^ Though the NPV for the secondary rule-out criterion (i.e. low risk strata) in our study was lower at 98.8%, this is in part attributable to our use of all-cause death as an outcome. For example, in a randomized controlled trial of the HIGH-STEACS pathway (HISTORIC trial), less than 25% of the composite 30-day outcomes in the rule-out group were attributable to cardiac death without MI, whereas in our study over 50% were due to all-cause death without MI, suggesting that up to one half of our death outcomes (and one quarter of our composite outcomes) were non-cardiac in nature. Furthermore, when considering patients with clinical concern for possible NSTE-ACS (as opposed to the broader umbrella of chest pain complaints), the true NPV for the secondary rule-out criterion is arguably better approximated by the sensitivity analysis of encounters with RISTRA eCDS use (99.7%).

### Subgroup analysis of rule-out criteria

We found that advanced age (≥ 65 years), known ischemic CAD (history of MI or coronary revascularization) and CKD (either stage 3 or stage 4+) resulted in significantly lower NPV for 30-day MI or death among encounters meeting rule-out criteria (point estimates of 99.2%, 98.6%, 98.9%, and 96.1%, respectively). On multivariable analysis, age ≥ 65 was the strongest independent factor, which may reflect inclusion of non-cardiac death in our composite outcome. However, age and known ischemic CAD subgroups were also investigated in the original HIGH-STEACS validation study using a composite outcome of 30-day MI or cardiac death with results nearly identical to our study (NPV for age > 65 of 98.9%; NPV for ischemic CAD 98.7%).^7^ Comparably diminished NPVs in the setting of advanced age or ischemic CAD have also been observed with the various ESC algorithms, including the 0/1-hour protocol^16, 23–25^, 0/2-hour protocol (using the Access hsTnI assay)^15^, and the older 0/3-hour protocol.^7^ A lower rule-out NPV in the presence of CKD, on the other hand, has not been as widely emphasized, with most investigations highlighting the limited PPV of elevated hs-cTn values in the presence of CKD, which we likewise observed.^26–30^ In all the forementioned instances, lower specificities and higher outcome incidences were the principal driving factors leading to decreased NPVs.

Clinical awareness of these limitations is warranted, with the following caveats: 1) applying additional clinical exclusions to achieve the commonly cited benchmark of 1% or less risk for adverse cardiac events may be “a bridge too far” owing to significant losses in efficacy (i.e. fewer patients ruled-out) with only marginal improvements in safety^31–33^ and, 2) the pre-test probability threshold for functional cardiac testing in the evaluation of possible NSTE-ACS may be as high as 2%.^34^

### Rule-in criteria performance

The KPNC NSTE-ACS algorithm rule-in criteria (derived from the 0/2-hour ESC protocol) also proved effective with a PPV of 60%, mirroring the results of a recent evaluation of the 0/2-hour ESC protocol using the Access hsTnI assay in a separate US population.^15^ As mentioned above, rule-in criteria PPV was reduced in the setting of CKD^26–30^ as well as in the setting of known CAD (nonischemic more than ischemic). Notably, younger age (<45) had the lowest associated PPV, a finding also seen when applying the ESC 0/1-hour protocol using a hs-cTnT assay, perhaps reflecting a higher relative risk for non-ACS causes of hs-cTn elevations (e.g. from pulmonary embolism) and lower risk of death in younger populations.^24^

### Clinical practice performance (ED disposition)

Considering the heterogenous performance of the rule-out criteria within clinical subgroups, it is reassuring that, in community ED practice, emergency physicians were able to select patients for discharge with improved safety compared to strict rule-out criteria application and with less loss of efficacy as compared to superimposed clinical risk scores (e.g. HEART score).^32^ Similarly efficacious improvements in safety were noted in a “real-world” study of the ESC 0/1-hour algorithm.^2^ It is further reassuring that this parsing was similarly effective within the higher risk subgroups of rule-out encounters. Identifying a structured means by which these gains can be reliably and consistently achieved without sacrificing efficiency remains an elusive end ^30, 32, 33^, though the application of machine learning technology appears promising.^35–37^

## Limitations

As a retrospective study, we were limited in our ability to identify patients with verified clinical concern for NSTE-ACS, particularly encounters without a chief complaint of chest pain or discomfort, and thus our findings may not be applicable to alternative presentations of ACS (e.g. isolated dyspnea). The use of diagnostic codes to adjudicate MI outcomes is also a limitation, though agreement has been shown to be good compared to prospective outcome adjudication^38^ and we were able to perform targeted manual chart review to improve capture of type 1 MIs following rule-out encounters that may have been miscoded as ACS, type 2 MIs or unstable angina, thus further bolstering the safety assessment of the algorithm. The exclusion of patients without active health plan insurance (to maximize outcome capture) may also limit the generalizability of our findings. Longer outcome follow-up periods may provide further insight, though the examined 30-day outcome window is more pertinent to ED disposition decisions and tends to correlate well with 1 year outcomes, especially among rule-out encounters.^10^

## Conclusion

The KPNC NSTE-ACS algorithm demonstrated excellent overall performance using the Access hsTnI assay when applied to ED patients across a large, integrated community health system in the US. Though rule-out criteria performance was modestly diminished in the presence of known ischemic CAD or advanced CKD, consistent with other reports, emergency physicians effectively mitigated these risks in the process of disposition decision making, providing reassurance that the “real world” algorithm performance conforms to current standards of care.

## Data Availability

Requests to access the data set from qualified researchers trained in human subject confidentiality protocols may be sent to KPNC at.

## Acknowledgements

Adina Rauchwerger, MPH for invaluable assistance with project management and support; the physicians of The Permanente Medical Group

